# MICRO-CT Imaging of Thiel-Embalmed and Iodine-Stained Human Temporal Bone for 3D Modeling

**DOI:** 10.1101/2020.12.15.20247536

**Authors:** Halm Sebastian, Haberthuer David, Eppler Elisabeth, Djonov Valentin, Arnold Andreas

## Abstract

**OBJECTIVE:** The following study investigates whether human Thiel-embalmed temporal bones are suitable for generating an accurate and complete data set with micro-computed tomography (micro-CT) and the effect of solid iodine staining for improved visualization and facilitated segmentation of middle ear structures.

**METHODS:** One temporal bone was used to verify the accuracy of the imaging by measuring the stapes digitally on the tomography images and physically under the microscope after removal from the temporal bone. All measurements were compared with literature values.

The contralateral temporal bone was used to evaluate segmentation and 3D modeling after iodine staining and micro-CT scanning.

**RESULTS:** The digital and physical stapes measurements differ up to 0.17 mm and 24% but correlate well with the literature values. Soft tissue structures were visible in the unstained scan. However, iodine staining increased the contrast-to-noise ratio by a factor 3.7 on average. The 3D model depicts all ossicles and soft tissue structures in detail, including the chorda tympani, which was not visible in the unstained scan.

**CONCLUSION:** Micro-CT imaging of Thiel-embalmed temporal bones accurately represents the entire anatomy. Iodine staining considerably increases the contrast of soft tissues, simplifies segmentation and enables detailed 3D modeling of the middle ear, which is suitable for further use as a finite element model (FEM).

## 1. INTRODUCTION

Virtual three-dimensional (3D) models can serve as a basis for computed dynamic models such as finite-element modeling (FEM). High resolution and high precision morphologic data are necessary for visualization and virtual 3D reconstruction of fine structures, as present in the middle ear. If the integrity of tissue is to be preserved, a widely used method is micro-computed tomography (micro-CT) imaging,^1-5^ since spatial resolution in the low micrometer to high nanometer range is possible. However, one drawback in micro-CT is the low visibility of soft tissue due to its weak absorption of x-rays. To improve the radiographic visibility and enable precise segmentation, the use of a contrast agent is desirable. Iodine solution (I_2_KI) has become a widely used contrast agents and has been reported to significantly improve the contrast-to-noise ratio (CNR) of submillimeter middle ear connective tissue structures.^6,7^ One drawback of iodine solution used for morphological imaging is the possibility of tissue shrinkage.^8^ Boyde et al. successfully applied solid iodine for vapor staining in order to avoid tissue shrinkage.^9^

Using fixated tissue is safer in terms of infectivity and tissue is longer usable for experiments. The fixation according to Walter Thiel has the further advantage of maintaining tissue suppleness, which is necessary for physiologic measurements (e.g. middle ear mechanics).^10^

The aim of this study is to investigate whether we can create a detailed and precise 3D model of a human middle ear by using solid iodine-stained Thiel-embalmed tissue and micro-CT imaging.

The resulting 3D model shall later on serve as future basis of a dynamic middle ear model using FEM, that will be validated and adjusted using physiological and pathophysiological motion data from laser doppler vibrometry (LDV) measurements of the same Thiel-embalmed temporal bone. The suitability of measuring ear micromechanics in Thiel-embalmed specimens using LDV has already been demonstrated.^11^

This study is divided into three separate parts:

- To determine whether Thiel-embalmed human temporal bone is suitable for micro-CT imaging.
- To evaluate the feasibility and contrast enhancement of iodine staining with regard to improved depiction of fine connective tissue structures (i.e. membranes, ligaments, tendons and muscles) within the tympanic cavity of a Thiel-embalmed specimen.
- To determine whether Thiel-embalmed human temporal bone micro-CT data allows the generation of an accurate and detailed 3D model of the human middle ear including delicate soft tissue structures. The accuracy of the 3D model is verified by measurement of different stapes dimensions.

## 2. MATERIAL AND METHODS

### 2.1 Thiel specimens

A male body donor in their 80’s was embalmed according to the Thiel method. ^10^ Two temporal bones with a sample size of 5 cm x 3 cm x 3 cm were excised. Two weeks prior to imaging, the temporal bones were removed from the Thiel solution and excess liquid was allowed to evaporate, since contrast differences are enhanced when samples are scanned in an air environment instead of liquid.

A micro-CT scan was acquired from the right temporal bone. This data set was imported in a 3D Slicer (Version 4.8.1, http://www.slicer.org) to measure the stapes dimensions.^12^ The stapes was then dissected from the temporal bone and measured under the microscope.

The second, left temporal bone was CT-scanned in its native state, then stained and re-scanned. The stained dataset was compared with the unstained and used to create the 3D model in the 3D Slicer. The detailed procedures are explained below.

### 2.2 Micro-CT imaging

Tomography scans were performed in a SkyScan 1172 micro-CT system (Bruker, Kontich, Belgium). Imaging conditions were a source voltage of 89 kV and a source current of 112 μA.

The x-ray spectrum was filtered with a 0.5 mm Aluminum/0.08 mm Copper filter. We used a 2 x 2 binned camera window with two overlapping lateral acquisitions (resulting projection size: 1336 x 3968 pixel) and acquired one projection image at every 0.2° degree of rotation resulting in 1018 projections. Each projection was exposed for approximately 2.6 seconds; three exposures were averaged into one to increase the signal-to-noise ratio. The resulting isometric voxel size (side length) was 13.27 μm. Projections were reconstructed into a set of PNG files using Nrecon (Version 1.7.0.4, Bruker) with a ring artifact correction of 16 and a beam hardening correction of 60%.

### 2.3 Sample preparation & iodine staining

In order to obtain a complete staining of the middle ear structures, iodine pellets were placed into the tympanic cavity after removal of some mastoid cells with a Luer forceps and a preparation needle. Access and integrity of the preserved structures (malleo-incudal joint, tendons and ligaments) were verified using the flexible Atmos FESS Portable 3.8 mm otoendoscope. The temporal bone (weight 22.6 g) and 0.4 g of solid iodine were put in a closed glass jar for 72 h. After iodine evaporation, the temporal bone was dark brown in color. Subsequently, the temporal bone was left to air dry at room temperature (average 21°C).

### 2.4 Unstained vs. iodine stained images

To evaluate theiodine staining effect, we compared the unstained and iodine-stained micro-CT data set. In particular, we assessed the following structures of interest (SOI): lateral malleal ligament, annular ligament, stapedius muscle, tensor tympani muscle and tympanic membrane.

The contrast of the SOI compared to air and bone was visually assessed and by calculating the CNR as follows: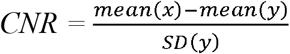.

We used Fiji (https://imagej.net/Fiji), an open-source platform for biological-image analysis^14^, for image processing and calculation of the mean pixel brightness values (x = pixel brightness of SOI, y = pixel brightness of background).

### 2.5 3D model

Using Fiji, we converted the .png files of the micro-CT scan into an .nrrd file, which was imported into the 3D Slicer. To enable optimal processing of the data set by the 3D Slicer, the dataset of the temporal bone was binned two-fold, resulting in images with a pixel size of 26 µm.

We segmented the middle ear structures from the dataset by manually painting the region of interest into each third slice, and used the “fill between the slices” function to interpolate in between the manually painted slices. The segmented dataset was pasted into a label map and further processed to yield a surface model.

### 2.6 Stapes measurements: 3D model vs. removed stapes

An important criterion when using micro-CT data set and 3D model for further computation is that Thiel fixation does not chemically impair middle ear structures and micro-CT imaging and segmentation within 3D Slicer represents the correct dimensions of the structures.

To verify this, we digitally and physically measured the stapes dimensions and compared them among each other and with literature values, respectively (Table 1). First, we measured the stapes on the image data set within the 3D Slicer (Figure 1). Thereafter, we dissected the stapes from the first temporal bone and measured the same dimensions under the microscope (Figure 2).

**TABLE 1.** STAPES DIMENSIONS AND DIFFERENCES

| <u>structure</u> | <u>dimension</u> |  |  | <u>differences</u> |  |  |
| --- | --- | --- | --- | --- | --- | --- |
|  | <u>3D Slicer</u> | <u>microscope</u> | <u>literature</u> | <u>Δ 1</u> | <u>Δ 2</u> | <u>Δ 3</u> |
| length footplate | 2.95 | 3.12 | $1.92^{18} - 3.56^{19}$ | 0.17 | (5) | |
| width footplate | 1.44 | 1.47 | $1.16^{20} - 1.4^{17}$ | | | 0.04/0.07 |
| height of footplate | 0.18 | 0.19 | $0.18^{18} - 0.39^{18}$ | | | |
| height of stapes | 3.36 | 3.49 | $3.07^{20} - 4^{17}$ | 0.13 | (4) | |
| length of stapes head | 0.98 | 1.04 | $0.85^{18} - 1.49^{21}$ | (0.06) | 6 | |
| width of stapes head | 0.74 | 0.75 | $0.65^{21} - 1.08^{21}$ | | | |
| width of anterior crus | 0.52 | 0.42 | $0.21^{18} - 0.65^{21}$ | (0.1) | 24 | |
| width of posterior crus | 0.43 | 0.41 | $0.14^{18} - 0.75^{21}$ | | | |
| area footplate | 3.15 | 3.6 | $2.7^{20} - 3.36^{20}$ | 0.45 | 12 | 0.24 |

*Note: Dimensions measured digitally (3D Slicer), physically (microscope) and range of literature values. Δ 1 and Δ 2: difference between our measuring methods in mm and % respectively. Δ 3 difference of our values to literature values. Values indicated in mm; Δ 2 in %; area in mm<sup>2</sup>.*

**FIG. 1:**
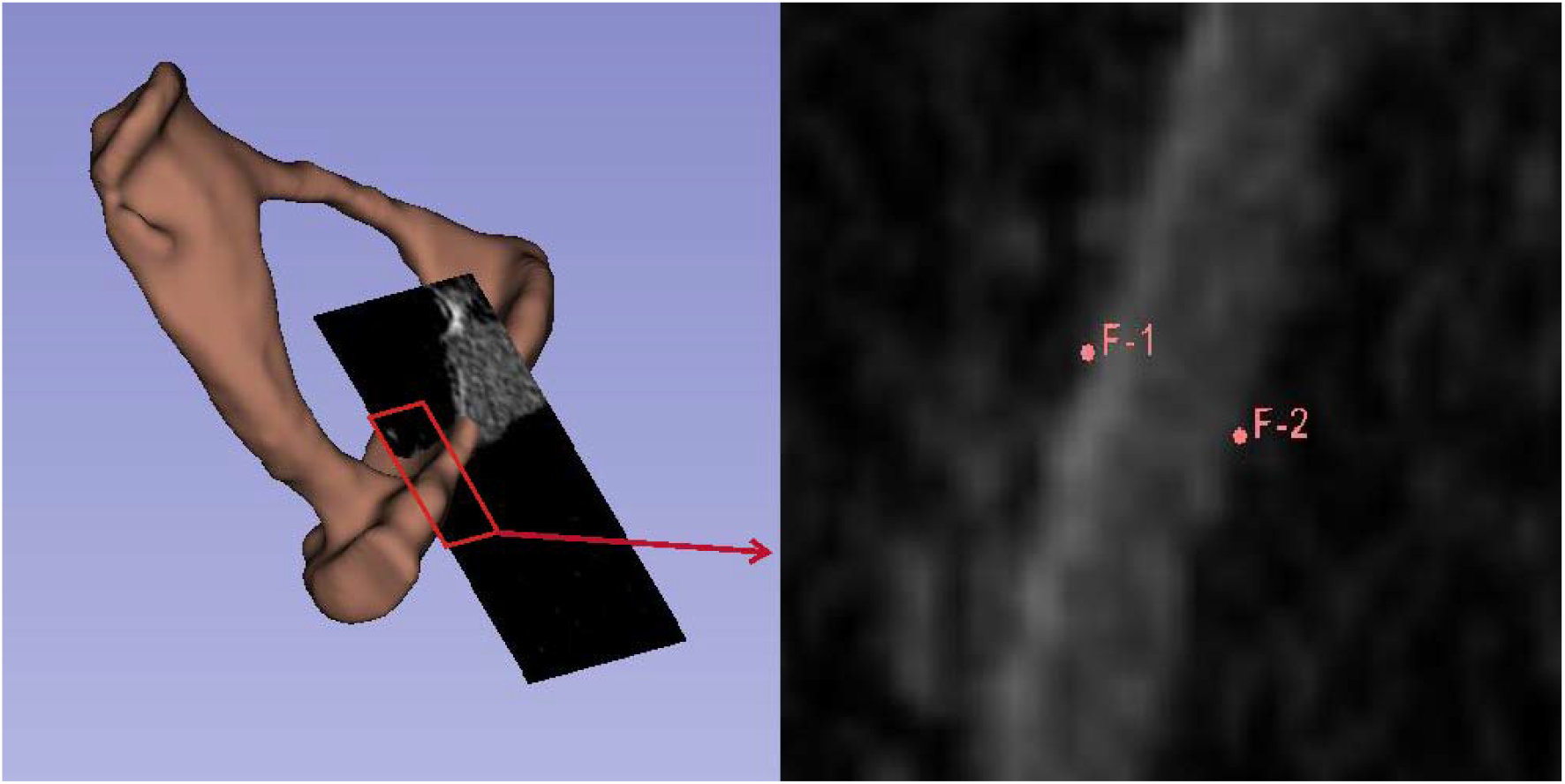
DIGITAL STAPES MEASUREMENT *Example for measuring stapes dimensions using 3D Slicer (height of the footplate). Left: 2D plane and created model viewed simultaneously. Right (zoom from left plane): placement of the measuring points*.

**FIG. 2:**
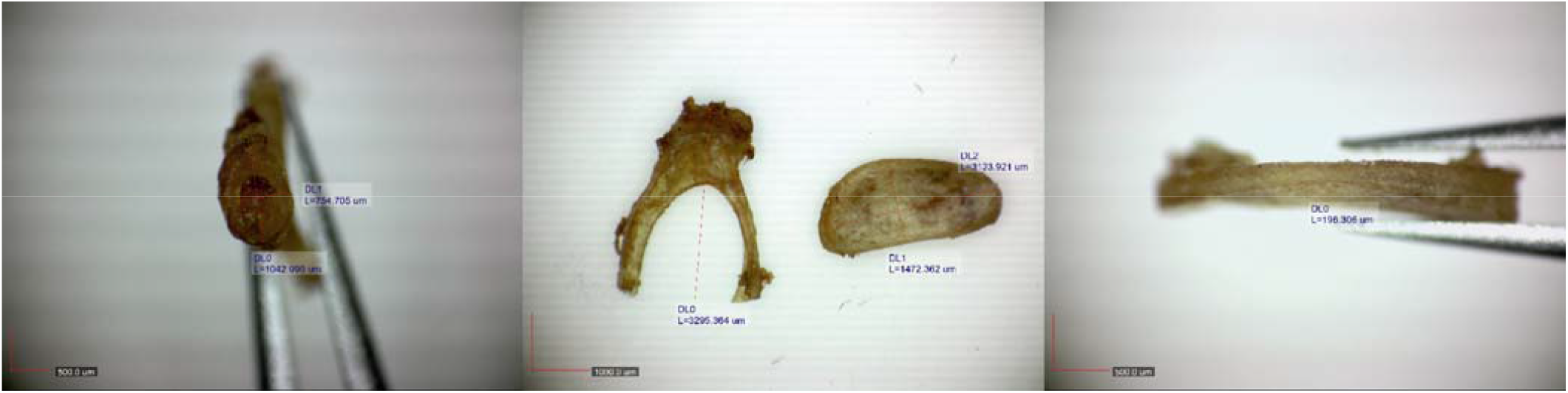
PHYSICAL STAPES MEASUREMENT *Measurement of the removed stapes with Dino Capture. Dimensions are indicated in Tab. 1*.

The stapes was photographed and measured with a light microscope Olympus Sz 40 (Olympus K.K., Shinjuku, Tokyo, Japan) using an AM7025X Dino-Eye Edge Eyepiece Camera (Anmo Electronics Corporation, New Taipei City, Taiwan). Camera resolution of 2592 x 1944 pixel results in an image pixel size of 6 µm. Images were processed using the Dino Capture 2.0 Microscope Imaging Software (Anmo Electronics Corporation). Special care was taken to measure the stapes in a comparable manner to the published literature values. A list with the exact measuring points is shown in Additional Table 1.

## 3. RESULTS

### 3.1 Unstained vs. iodine stained images (Figure 3)

Most of the middle ear soft tissue structures were visible with native micro-CT imaging. Notably, the annular ligament was identified as a gap between the stapes footplate and the adjacent promontory bone. Interestingly, after iodine staining, the annular ligament was depicted with a contrast similar to the bone directly adjacent to it, which made it more difficult to recognize than without iodine staining (Figure 3A).

**FIG. 3.**
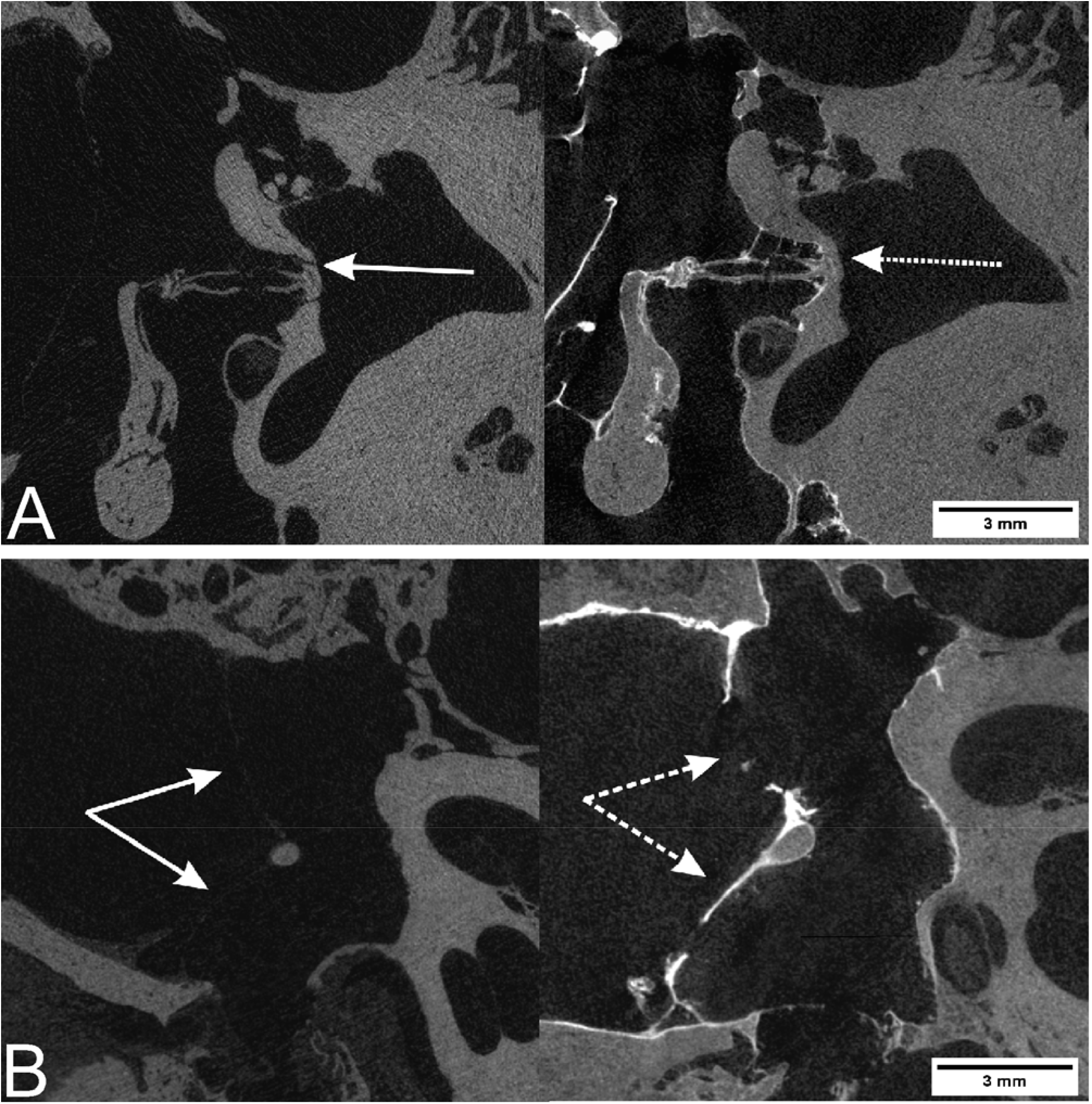
A,B: INFLUENCE OF IODINE STAINING ON MICRO-CT IMAGING *The continuous arrow represents the unstained structure, the dotted arrow the iodine stained structure. A: Annular ligament. Note: the ligament is better visible in the unstained sample. B: Tympanic membrane. Note: unstained area in the lower and perforated area in the upper part (probably retraction after staining)*.

**FIG. 3.**
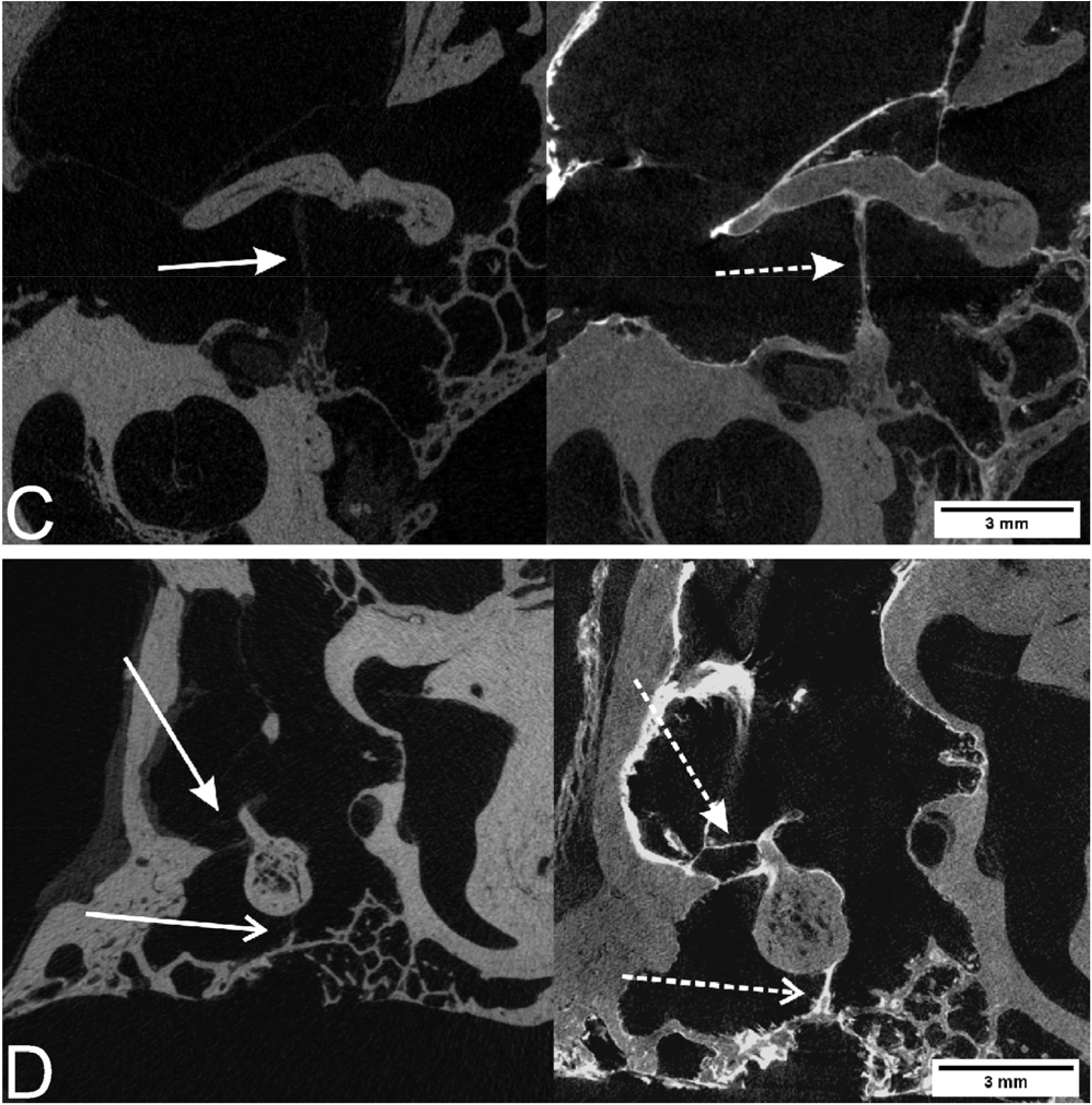
C,D: INFLUENCE OF IODINE STAINING ON MICRO-CT IMAGING *The continuous arrow represents the unstained structure, the dotted arrow the iodine stained structure. C: Tensor tympani muscle. D: Ligaments of malleus. Open arrowhead = superior malleal ligament. Filled arrowhead = lateral malleal ligament*.

Thus, the images acquired after iodine staining did not show any previously occult structures, except for the chorda tympani (explanation below).

However, it should be emphasized that visibility and demarcation of the SOI were pronouncedly enhanced after staining, due to an improved contrast of the soft tissue to air and bone (Figures 3). The CNR values of the SOI were, on average, 3.7 times higher (Range: 1.17 – 5.59, SD: 1.83) than the CNR of the unstained soft tissues (Figure 4). A detailed presentation of the calculated CNR values is shown in Additional Table 2.

**FIG. 4:**
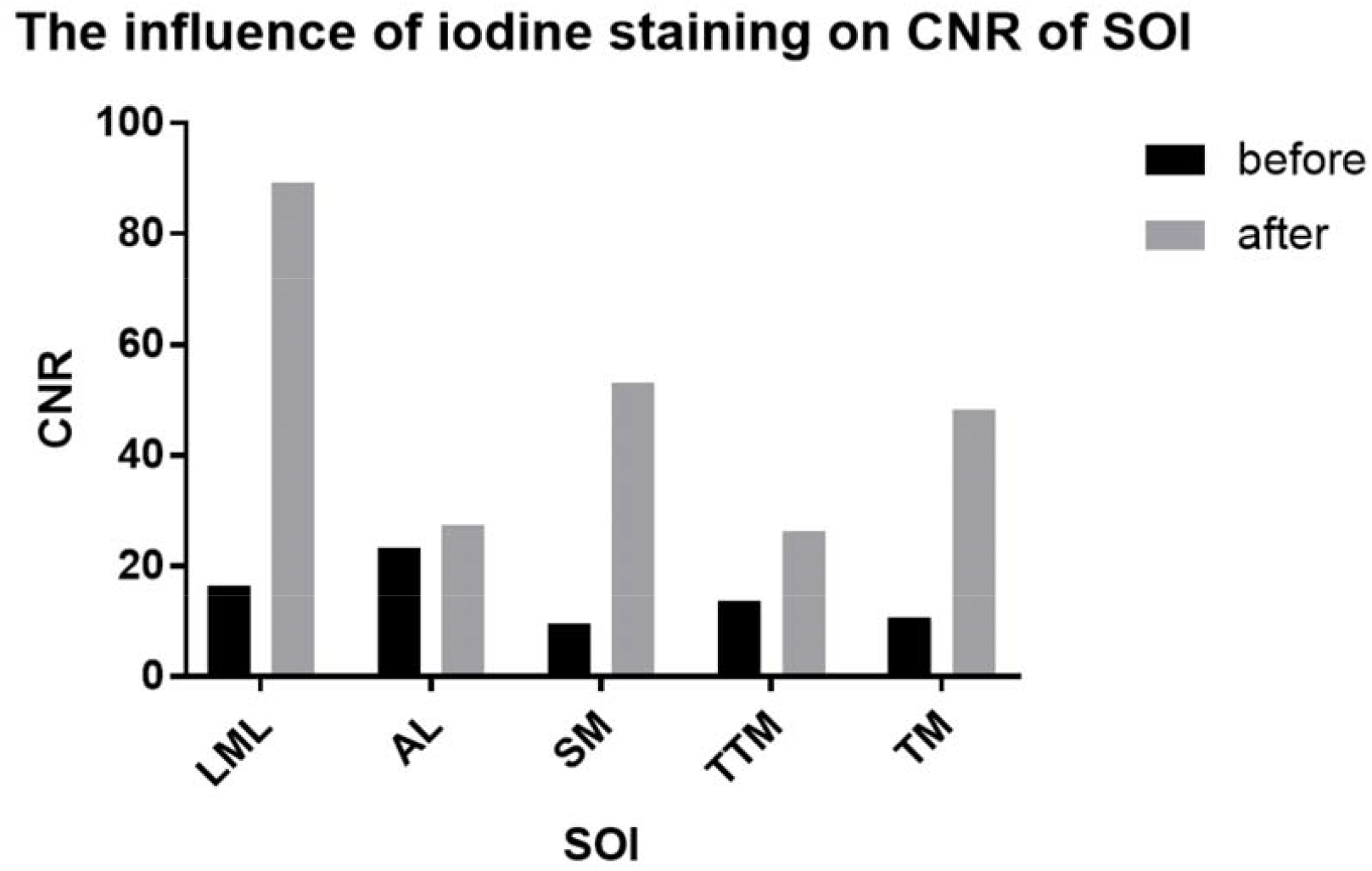
STAINING EFFECT ON CNR *Contrast to noise ratio (CNR) before and after staining. SOI = structures of interest. LML = lateral malleal ligament. AL = annular ligament. SM = stapedius muscle. TTM = tensor tympani muscle. TM = tympanic membrane*.

### 3.2 3D model

The improved visibility due to the staining has led to a better delimitation from the background and better assignment of middle ear soft tissue structures (except for the annular ligament, despite a slightly higher CNR). This made segmentation considerably easier and more accurate. The small perforation of the tympanic membrane, visible by micro-CT (Figure 3B), was manually closed during the segmentation procedure. It is remarkable that even the chorda tympani, which was invisible in the unstained scan, was distinguishable. Figure 5 shows the course of the chorda tympani between malleus and incus.

**FIG. 5:**
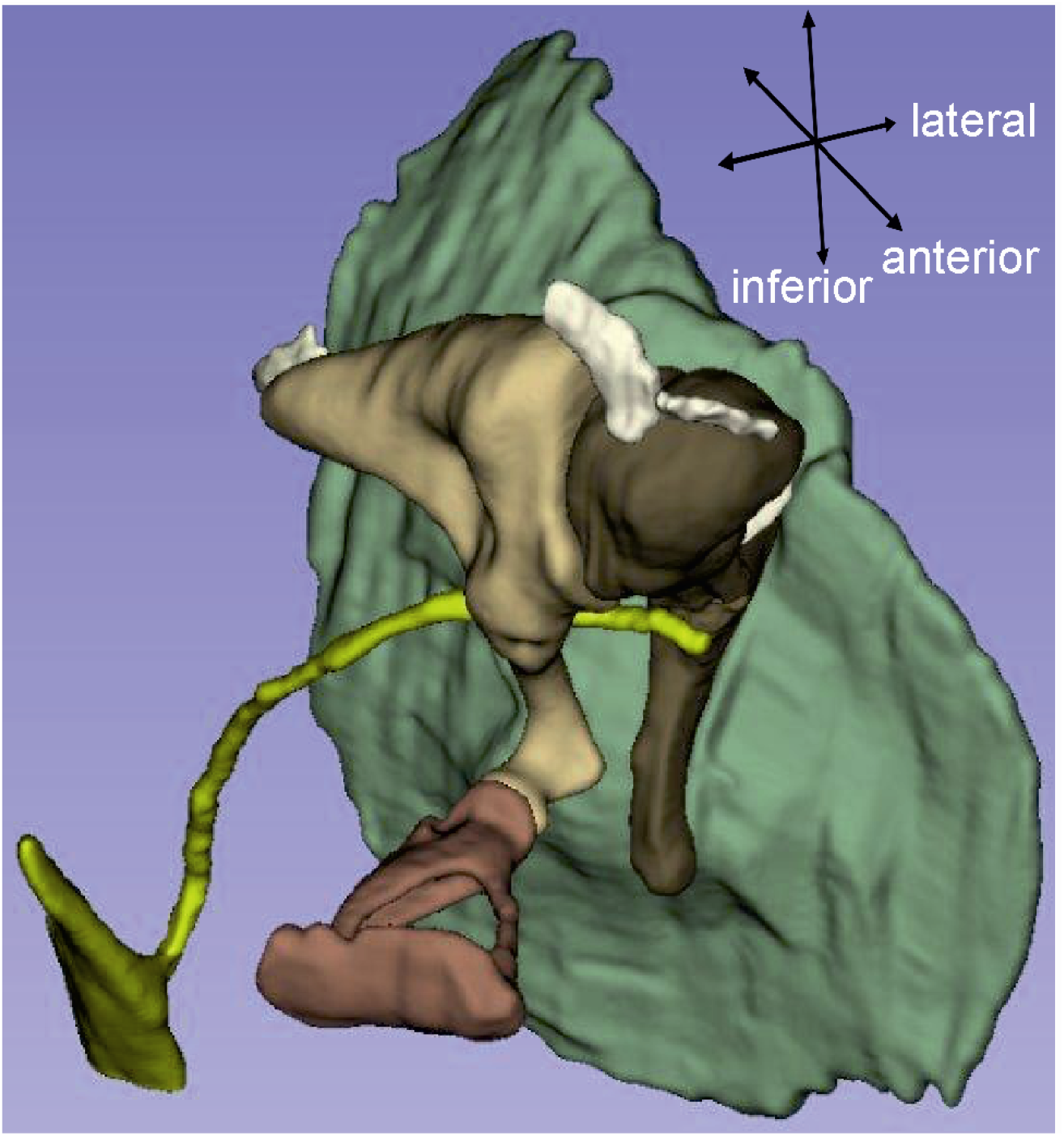
3D MODEL *3D model of the middle ear structures segmented in 3D Slicer*.

The resulting 3D model accurately depicts the 3D arrangement and position of the middle ear bones, as well as the detailed position, insertions and tensile direction of the associated soft tissues.

### 3.3 Stapes measurements: 3D model vs removed stapes

Table 1 shows the digitally and physically measured stapes dimensions, literature references and the three largest (absolute and percentage) measurement deviations.

High percentage deviations occurred rather in small stapes structures and high absolute deviations rather in large stapes structures. For tiny structures, small absolute deviations already caused large percentage differences. In general, the measured values of both methods differed by 0.01 – 0.17 mm or 1 – 24%, respectively.

### 3.4 Stapes measurements: 3D model & removed stapes vs literature values

The width of the footplate of 1.44 mm (3D Slicer) and 1.47 mm (manually), respectively, are about 40 µm and 70 µm wider than the largest reference value in the literature.

The area of the footplate is about 0.24 mm ^2^ larger than the largest reference values in the literature. Six out of nine (66%) of the physically measured values and eight out of nine (88%) of the digitally measured values were within a range of ± 1 SD of the mean as compared to stapes measurement values in the literature (Additional Table 3).

## 4. DISCUSSION

In this study, we demonstrate that a high-resolution micro-CT dataset could be obtained of a Thiel-embalmed human temporal bone. The resulting images depicted all structures of interest including fine connective tissue structures, which were even more enhanced (augmented CNR) after application of iodine vapor staining. Furthermore we were able to segment the image data set and thus generate a detailed 3D model of the human middle ear including soft tissue structures. Using stapes dimensions, the generated 3D model corresponds well when compared to the physically measured ossicle as well as data from the literature.

### 4.1 Unstained vs. iodine stained images

Micro-CT imaging of unstained Thiel-tissue provided image data in high detail and all middle ear structures, except the chorda tympani, were recognizable (Figures 3).

However, to improve the visibility and above all to visualize the trajectory of small connective tissue structures for facilitated and precise segmentation, staining is still desirable. An improved contrast enhancement and tissue depiction by iodine staining was shown by Metscher,^15^ while Rohani reported a quantitative improvement of the CNR.^7^

By using solid iodine, Boyde and co-workers avoided possible tissue shrinkage when staining with iodine solutions.^8,9^ Analogous to this procedure, we used solid iodine for better contrast and placed it directly into the middle ear after removing some mastoid cells.

To the best of our knowledge, the effect of vapor iodine staining on Thiel embalmed specimen has not been reported in the literature to date. The iodine dose used in this study was empirically determined according to experiments with zebrafish (unpublished data from our institute), which based on a work of Babaei.^16^ We used about 17 mg of solid iodine per gram of sample, staining for 72 h, which showed good results.

For comparison, Boyde and co-workers used between 8-125 mg solid iodine per gram sample and achieved best results when staining for up to 7 days.^9^ In our study, solid iodine staining of the temporal bone specimen clearly enhanced the visibility of middle ear soft tissue structures (Figures 3) and improved the CNR of the SOI by a factor up to 5.59 (mean 3.7, SD 1.83; Figure 4). Analysis of micro-CT images and the anatomical orientation definitely benefits from iodine staining.

### 4.2 3D Model

Increased contrast is a notable advantage for analyzing the micro-CT image set resulting in an easier, thus quicker, but also more accurate segmentation. In contrast to our expectations even the chorda tympani was visible and could be integrated into the 3D model.

As a notable exception, the annular ligament showed no improvement in detectability as the iodine staining made the contrast similar to the surrounding bone.

Segmentation resulted in a precise 3D model, which showed in detail the complex middle ear anatomy (Figure 5). Therefore, our 3D model may be a suitable base for further use in FEM.

### 4.3 Stapes measurements: 3D model vs. removed stapes

The difference between both measuring methods was up to 170 µm or 24 %, depending on the measured stapes structure (Table 1).

A possible explanation is that the corresponding structure had a slightly different orientation when measured digitally and physically. Furthermore, measurement differences may occur due to measuring inaccuracy related to the pixel size. A difference of 170 µm with a pixel size of 26 µm means a deviation of the measurement point of only 6.5 pixels (3D Slicer) and 28 pixels with a pixel size of 6 µm (Dino Lite camera). When a higher magnification of an image is selected to set accurately the measurement point, the individual pixels become visible. The main difficulty about setting the measurement point, is the image transition from bone to air.

At high magnification, there is no clear cut-off. The border of the bony subject fades out with a gradual transition of pixels brightness from bone to air, posing a source of inaccuracy for measurement point setting (Figure 1). For small pixel size (6 µm), there are more pixels to choose from but the resulting deviation is small, because the pixel size is small. For a larger pixel size (26 µm), there are fewer pixels, but the resulting deviation is greater.

### 4.4 Stapes measurements: 3D model & removed stapes vs literature values

All values correspond well with the literature values (Table 1). Nonetheless, two dimensions must be explained.

First, the manually measured width of the stapes was 1.47 mm and the 3D Slicer-measured width 1.44 mm. These values moderately deviate from the 1.4 mm given in the literature,^17^ which, however, is only reported to one decimal place.

Secondly, the manually measured area of the stapes footplate was 3.60 mm^2^, which is about 0.24 mm^2^ larger than the largest reported value.

However, when calculating the area using the largest reported width and length, the resulting footplate area would be 4.11 mm^2^ and consequently our measured area within the range. Overall, our measured values correspond well with the literature values.

Therefore, we consider the chemical influence of the Thiel fixation on the tissue as negligible and micro-CT imaging of Thiel-fixated tissue as a precise representation of the anatomy.

If anatomical structures in general should be measured, the following advantages and disadvantages can be mentioned about digitally measurements on a 3D model like in 3D Slicer. Advantages are:

- It is a nondestructive procedure, and
- It allows a simultaneous view of the three image planes and the 3D model for better setting of the measurement points while avoiding projection error (Figure 1).

The disadvantages are:

- The process of image acquisition and precise segmentation is time-consuming. The time spent on segmentation greatly influences the accuracy of the 3D model, and
- Binning may be necessary, depending on the size of the image data set and the computer memory.

### 4.5 Limitations

The fixation protocol can be adapted by anatomical institutes. Depending on changes, this may have an impact on the tissue and require a re-validation of the described method.

The sample size of two temporal bones may mean that certain confounders have not been detected. Although we do not believe in a fundamental change in the statements of the present study further studies are needed to support our findings.

Furthermore, the study did not verify the effect of tissue shrinkage after application of solid iodine.^9^

Thiel embalming requires many chemicals and is time-consuming. The costs are described as 8-14 x higher than formalin fixation (300-437 Euro vs. 30-36 Euro).^22, 23^ It must be noted that Thiel specimen are validated and used for the study of middle ear mechanics^11^, which is not possible with formalin specimen due to tissue stiffness. Cost-reducing measures (storage of several cadavers in one tank; multiple use of a cadaver for different clinical courses and experiments) are not yet considered. The cost for Iodine is negligible. 3D Slicer and Image J are freeware.

## 5. CONCLUSION

Thiel-embalmed temporal bones are suitable for detailed presentation of the anatomy using micro-CT imaging. Iodine-vapor-staining substantially improves the contrast of soft tissue structures in micro-CT imaging. The considerably simplified segmentation of all the structures allows the generation of an accurate 3D model.

We conclude that the use of Thiel-fixated tissue, vapor iodine staining and micro-CT imaging is a suitable method to create a 3D model, which can further be used for FEM computing.

## 6. DECLARATIONS

### 6.1 Ethics approval and consent to participate

Human cadaveric material was used according to the current Swiss Federal Act on Research involving Human Beings (Human Research Act, HRA) (Federal Act on Research involving Human Beings) and the current guidelines of the Swiss Academy of Medical Sciences (https://www.samw.ch). Donors formally consent to the post-mortem use of body parts for research purposes. Approval by swissethics was granted (PB_2018-00113).

### 6.2 Availability of data and materials

All data generated during this study are included in this published article and as additional data files.

## 6.5 Acknowledgments

Many thanks to Marek Kaminek, Institute of Anatomy, University of Bern, who operated the Dino Lite camera, photographed and measured the stapes.

## 8. LEGENDS

### 8.1 Additional Data Files

ADDITIONAL TABLE 1: Detailed position of the measuring points used.

ADDITIONAL TABLE 2: Pixel brightness values of the structure of interest (SOI) and the noise, each for the unstained and stained micro-CT image, respectively. CNR formula see section 2.4.

ADDITIONAL TABLE 3: List of stapes dimensions found in literature compared with our digital (3D Slicer) and physical (microscope) measurements. Values in mm. Green color indicates our own values within the SD of the literature values. Orange color indicates the ratio of our values within the SD and of the literature values.

